# Upper arm length along with mid-upper arm circumference to enhance wasting prevalence estimation and diagnosis: sensitivity and specificity in 6 to 59 month-old children

**DOI:** 10.1101/2020.05.12.20089433

**Authors:** Mouhamed Barro, Mohamed Daouda Baro, Djibril Cisse, Noel Zagre, Thierno Ba, Shanti Neff-Baro, Yacouba Diagana

**Author notes:** **Corresponding author:** Mouhamed Barro, 2 Square de la poterne 91300 Massy.

## Abstract

**Objective:** To evaluate the added value of the use of upper arm length (UAL) along with MUAC (mid-upper arm circumference) to diagnose and estimate the prevalence of wasting in comparison to current WHO standard and others MUAC based methods.

**Design:** We included UAL to usual anthropometric measurements during a Mauritanian national 6-59-month-old cross-sectional nutritional survey. Children were classified into 3 groups UALG1, UALG2 and UALG3 according to the following UAL limits: ≤ 150 mm, 151-180, and > 180mm respectively. We used a Receiver Operating Characteristic curve to determine the best MUAC cut-off for each group with weight-for-height Z score as a reference standard. We compared the wasting prevalence, sensitivity, and specificity, of all diagnostic methods.

**Findings:** In total, 12 619 children were included in the study. Wasting prevalence was 16.1%, 5.0% and 12.5% when diagnosed by WHZ < −2, MUAC < 125 mm and MUAC-UALG methods respectively. Using the MUAC-UALG method increased the sensitivity for wasting diagnosis from 17.98 % with MUAC < 125 mm to 39.43% with MUAC-UALG. The specificity decreased from 97.49% with MUAC < 125 to 92.71% with MUAC-UALG. With MUAC-height Z score and MUAC < 138 mm, sensitivity was 26.04% and 69.76%and specificity were 97.40%and 75.64% respectively.

**Conclusion:** This alternative method using MUAC measuring tape to measure UAL increases the wasting diagnosis accuracy and allows for a better estimation of wasting prevalence. This method could be used as a potential alternative method for quick surveys in emergency settings such as Corona virus disease 2019 context.

## Introduction

Wasting is a major public health problem in low- and middle-income countries. The risk of death is higher in wasted children defined by a weight-for-height z-score (WHZ) below −2, when compared to non-wasted children^(1)^. When diagnosed with wasting, children can be treated at home^(2)^. The earlier the child is diagnosed the shorter the duration of the treatment.^3^ However, wasting screening and diagnosis has been a challenge for the entire humanitarian community. WHZ remains difficult to obtain routinely at the community level as it requires heavy equipment and well-trained staff. Mid-upper arm circumference (MUAC) is therefore preferred in the field due to its simplicity (MUAC < 115mm for severe wasting, MUAC < 125 mm for wasting) as per the WHO recommendations.^1^ However, MUAC has shown its limits for wasting diagnosis as well as prevalence estimation.

In 2019, wasting (as defined by WHZ score below −2) affected more than 47 million children under five years old world-wide.^4^ Although both low WHZ and MUAC are recommended for wasting diagnosis,^1^ only low WHZ is used for wasting prevalence evaluation by WHO.^4^ The use of current WHO’s MUAC cut-off recommendation does not allow for wasting prevalence estimation with an acceptable accuracy.^5^

Different MUAC cut-offs have been proposed in the past decades for wasting diagnosis (also called acute malnutrition). In the 1960’s, a study based on a population of non-malnourished Polish children showed that MUAC had little or no relation to age and gender in children aged one to five years^(6)^. Shakir A. and Morlaey D suggested a coloured cord to measure upper-arm circumference for screening and diagnosis of wasting in children 6-59 months-old^(7)^. Children were categorized in three groups according to their MUAC: red, yellow and green for MUAC under 125 mm, between 125 mm and 135 mm, and over 135 mm respectively. In 1985 Bernt Lindtjorn showed that these cut-off points greatly exaggerate wasting prevalence rates and proposed new cut-off points (110 mm and 130 mm)^(8)^. Benr and Nathanail compared the WHZ < −2 and MUAC <125 mm methods and concluded that these two methods identify similar proportions of wasted children^(9)^. However, beyond the cut-off itself, the use of a single cut-off for wasting diagnosis in all children within this age range has been debated^(10,11)^. Indeed, MUAC has been reported to be age specific and the use of MUAC with a single cut-off underestimates wasting in older children^(12,13)^. To address this bias, a MUAC-based method taking into account child’s age and sex has been implemented. A Z-score is assigned to each child according to their MUAC, age and sex^(14)^. However, the difficulty of determining the children’s age led to the use of another index, based on MUAC, height, and sex^(15)^. These methods certainly improve the sensitivity of wasting diagnosis but are not simple enough to be used for routine diagnoses. In fact, the determination of the children’s age on the one hand and their exact height on the other hand are essential for the MUAC-age and MUAC-height indices. Due to the necessity of calculating the Z-score for each child, both methods are not really routinely used in the field.

We therefore considered an alternative method of wasting diagnosis with greater sensitivity and greater potential for routine use. Children’s height or age is not required. The method is based on the use of MUAC in relation to child’s upper arm length (UAL) which can be measured at the same time as the MUAC measurement, using the same measuring tape. We tested this method in a nutritional survey conducted in July 2015 according to the methodology “Standardized Monitoring and Assessment of Relief and Transitions” (SMART) in Mauritania. The current study aimed at evaluating the added value of the use of UAL along with the MUAC to diagnose and estimate the prevalence of wasting in comparison to the WHO standard as well as other MUAC based methods.

## Methodology

### Data collection

Data collected from the national SMART survey conducted in Mauritania in 2015 were used for the present study^(16)^. It was a cross-sectional survey with two-stage random sampling, led by the nutrition department of the Ministry of Health with technical support from UNICEF. The survey followed SMART survey’s guideline^(17)^. All of the measurements were carried out by teams of trained investigators who were experienced in taking anthropometric measurements. A national representative sample of children under five years old was used for this survey.

Weight was measured with a precision of 100 g using an electronic SECA type weighing scale. Height was measured in cm with a precision of 0.1 cm using SHORR toises. MUAC was collected in all children aged 6 to 59 months with precision to 1 mm using MUAC tapes. UAL was measured by the same MUAC tape as those used for MUAC measurement. This length corresponds to that used to determine the mid-upper arm location, namely the length between the tip of the elbow (the olecranon) and the tip of the scapula (acromion). The oedema was systematically searched at the top of both feet by exerting a pressure with the thumb for 3 seconds. Standardization of the measurements and plausibility checks were done according to the standards and recommendations of the SMART methodology^(17)^.

### Data analysis

After a double entry to clean the anthropometric data, Z-scores were calculated using ENA Delta software November 2014. Children were excluded from the analysis based on the following criteria: MUAC, height, sex or weight not recorded, extreme WHZ (< −5 or > +5), or arbitrarily considered extreme UAL (< 7 cm or > 30 cm). Wasting by low WHZ was defined by (WHZ < −2 using the 2006 WHO growth reference. Wasting by low MUAC-height Z score (MUAC-HZ) was defined by MUAC-HZ < −2. Wasting by MUAC-125 mm (MUAC-125) was defined by MUAC < 125 mm. Additionally, we compared our diagnosis approach with another MUAC cut-off proposed by Laillou and colleagues, wasting by MUAC 138 mm (defined by MUAC < 138 mm)^(18)^. Wasted children (according to the WHZ <-2) were divided into the following three groups of the same size, according to their UAL: Children with UAL ≤ 150mm, 151 ≤ UAL ≤ 180mm and UAL ≥ 180mm were classified in UAL group1 (UALG1), UAL group2 (UALG2) and UAL group3 (UALG3) respectively (Figure 1). In order to diagnose wasting by MUAC with different cut-offs, three different cut-offs were established for each UALG. Receiver operating characteristic (ROC) methodology was used to determine new MUAC cut-offs with improved sensitivity for wasting diagnosis for each UALG with a minimum specificity of 90% (S1). Data were analysed using IBM SPSS statistics software.

**Figure 1:**
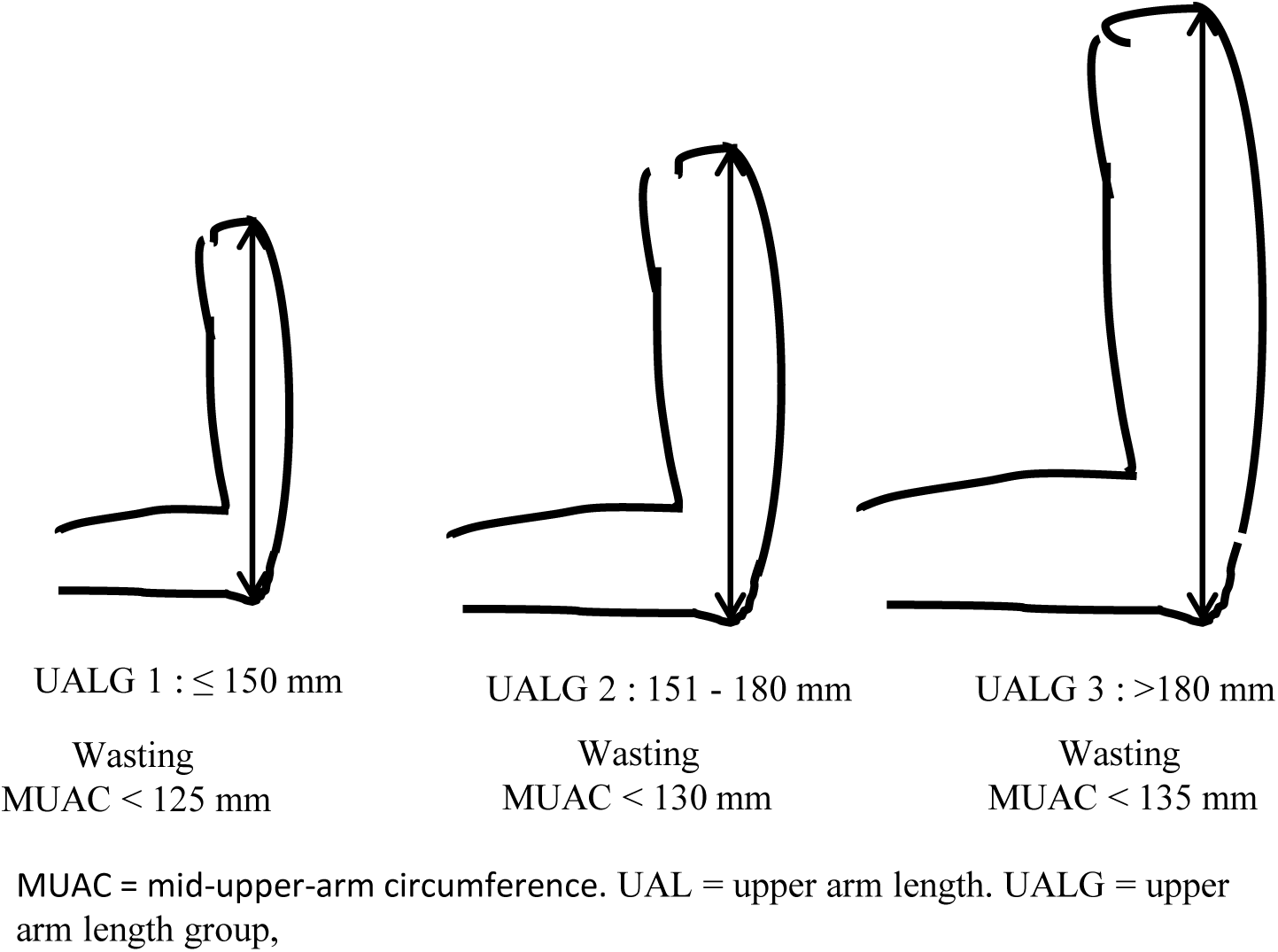
Classification of children according to their UAL and MUAC cut-off for each UALG.

The accuracy of our diagnosis method was evaluated according to the STARD recommendation^(19)^. Wasting by WHZ <-2 was used as reference standard to calculate the sensitivity and specificity of all the diagnosis methods that we tested. Sensitivity reflects the ability of the test to identify wasting among those identified by WHZ <-2. Specificity reflects the ability to correctly identify non wasted cases among those identified by WHZ >-2. Medcalc online version (https://www.medcalc.org/calc/diagnostic_test.php) was used to calculate sensitivity, specificity, positive predictive value, negative predictive value, with 95% confidence interval for each wasting diagnosis method.

### Statistic tests

Mean and standard deviation were calculated for continuous values. Correlations between continuous variables were evaluated using pearson test. Mean UAL, MUAC, age, height, and WHZ comparison among UAL groups was performed by Student T-tests. Wasting prevalence was calculated for each wasting diagnostic method.

## Results

Anthropometric measurements were taken from 12,626 children aged 6 to 59 months throughout Mauritania. In total, 36 children (<0.29%) presenting missing or inaccurate data were excluded from analysis (figure 2). A total of 12,590 children with 49.9% girls were included in this study. No child was found with bilateral oedema during the survey.

**Figure 2:**
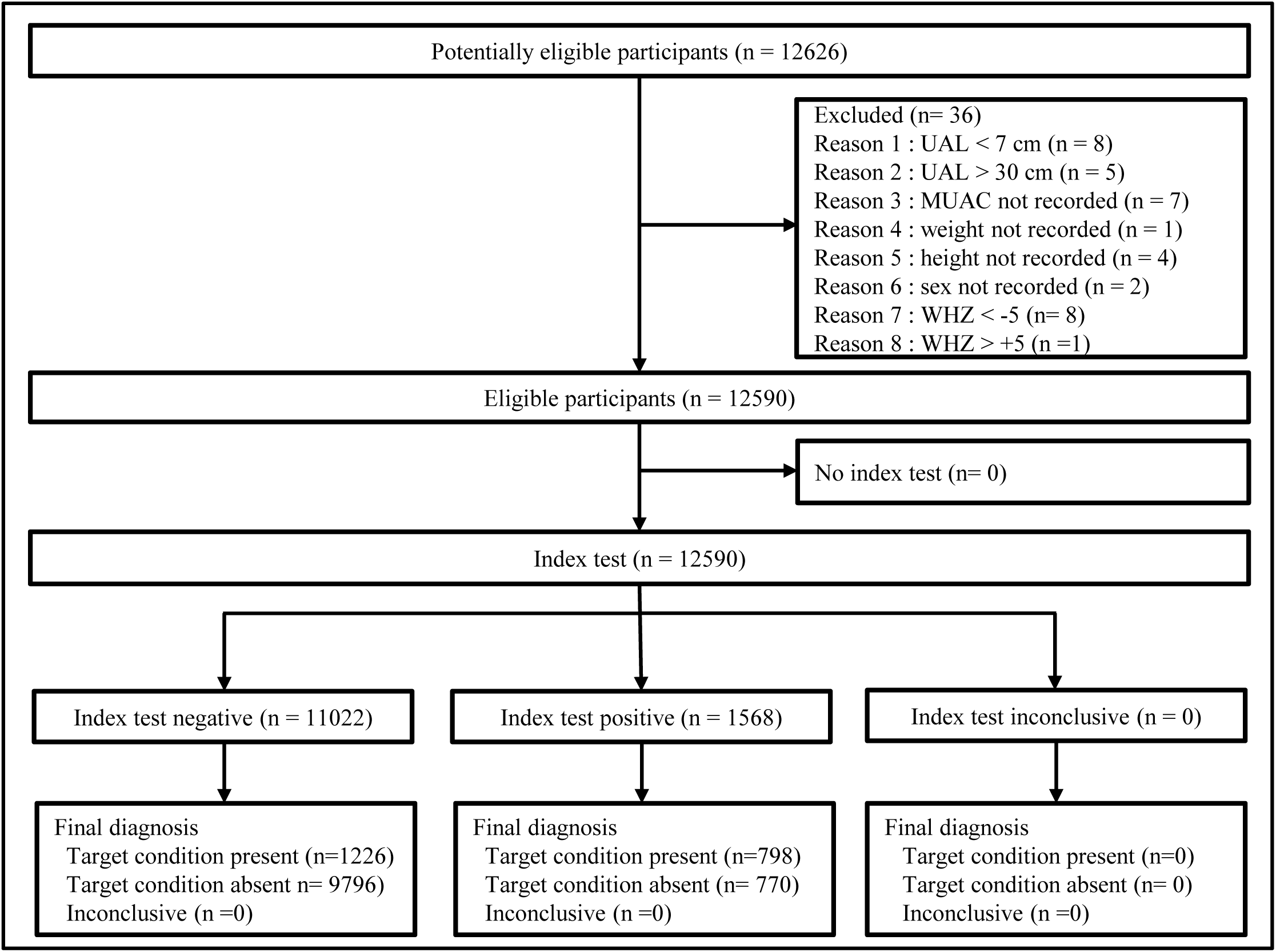
Flow of participants for wasting diagnosis test. Children with not recorded MUAC, weight, height, or sex were excluded. Children with too high or too low UAL were excluded. MUAC = mid-upper arm circumference, UAL = upper arm length, WHZ = weight-for-height Z score.

Our results demonstrated that UAL was correlated to height (pearson correlation = 0.65, p< 0·001) and age (pearson correlation = 0.62, p< 0.001) and MUAC was correlated to age (pearson correlation = 0.45, p< 0.001) as well as height (pearson correlation = 0.51, p< 0.001)).

Using ROC curves with WHZ as a reference standard allowed to determine the following MUAC cut-offs for each UALG for wasting diagnosis: 125mm, 130mm and 135mm for UALG1, UALG2 and UALG3 (figure 1 and S1).

The mean and standard deviation of childrens’ age, weight, and MUAC are described in table 1. Mean MUAC, height, and age significantly increased with UALG (p < 0.001) (table 1). The prevalence of wasting as determined by WHZ < −2 was 12.5% (table 2). When evaluated by MUAC-UALG method, wasting prevalence was 16.1%. With MUAC-125, MUAC-HZ and MUAC-138 the prevalence of wasting was 5.0%, 6.3% and 31.7% respectively.

**Table 1:** Anthropometric measurements by UALG. UALG1 ≤ 150mm, 150 ≤ UALG2 ≤ 180, UALG3 > 180 mm. T test was used to compare all continuous variables. The p values were < 0.001 between UALGs for all tested variables.

|  | n | Age,<br>months |  | Weight,<br>kg |  | Height, cm |  | MUAC, mm |  |
| --- | --- | --- | --- | --- | --- | --- | --- | --- | --- |
|  |  | Mean | SD | Mean | SD | Mean | SD | Mean | SD |
| UALG1 | 2582 | 17.9<sup>a</sup> | 12.2 | 8.8<sup>a</sup> | 2.4 | 75.6<sup>a</sup> | 10.3 | 137.3<sup>a</sup> | 12.1 |
| UALG2 | 4224 | 25.0<sup>b</sup> | 11.3 | 10.1<sup>b</sup> | 2.0 | 82.1<sup>b</sup> | 8.2 | 141.2<sup>b</sup> | 10.9 |
| UALG3 | 5784 | 39.4<sup>c</sup> | 13.0 | 12.6<sup>c</sup> | 2.4 | 92.9<sup>c</sup> | 9.4 | 147.3<sup>c</sup> | 11.3 |
| Total | 12590 | 30.2 | 15.2 | 11.0 | 2.8 | 85.7 | 11.6 | 143.2 | 12.0 |

MUAC: mid-upper-arm circumference; UALG: upper arm length group.

**Table 2:** Wasting prevalence determined by different methods.

|  | Wasting |  |  |  |  |
| --- | --- | --- | --- | --- | --- |
| Wasting indicators | WHZ < -2 | MUAC-125 | MUAC-138 | MUAC-HZ | MUAC-UALG |
| n | 12590 | 12590 | 12590 | 12511 | 12590 |
| Prevalence (%) | 2024 (16.1%) | 629 (5.0%) | 3986 (31.7%) | 794 (6.3%) | 1568 (12.5%) |

WHZ < -2: weight-for-height Zscore < -2; MUAC-125: mid-upper-arm circumference < 125 mm; MUAC-138: mid- upper-arm circumference < 138 mm; MUAC-HZ: MUAC-height Z score; MUAC-UALG: mid-upper-arm circumference per upper arm length group.

The diagnosis test accuracy for each indicator is summarised in the table 3. Overall, MUAC-125 had the lowest sensitivity (17.98% [16.33%; 19.73%]) and the highest specificity (97.49% [97.18; 97.78]) (table 3). With single fixed cut-off indicators (MUAC-125 or MUAC-138) sensitivity decreases, and specificity increases with UALG. This was not observed with adapted cut-offs (MUAC-HZ or MUAC-UALG) (S2). Although MUAC-138 had the highest sensitivity (69.76% [67.71; 71.76]), it had the lowest specificity (75.64% [74.81; 76.45]) leading to more than 24 % false positives. MUAC-UALG had a higher sensitivity (39.43% [37.29; 41.59]) than MUAC-125 and MUAC-HZ. MUAC-UALG had a higher specificity than MUAC-138 and a lower specificity than MUAC-HZ and MUAC-125.

**Table 3:** Wasting diagnosis accuracy based on sensitivity, specificity, positive and negative predictive value for each indicator. Weight-for-height z-score < −2 was used as reference standard.

|  | Sensitivity |  | Specificity |  | Positive predictive value |  | Negative predictive value |  |
| --- | --- | --- | --- | --- | --- | --- | --- | --- |
|  | % | 95% CI | % | 95% CI | % | 95% CI | % | 95% CI |
| Wasting by MUAC-125 | 17.98 | 16.33 , 19.73 | 97.49 | 97.18 , 97.78 | 57.87 | 54.15 , 61.50 | 86.12 | 85.87 , 86.37 |
| Wasting by MUAC-138 | 69.76 | 67.71 , 71.76 | 75.64 | 74.81 , 76.45 | 35.42 | 34.42 , 36.44 | 92.89 | 92.43 , 93.32 |
| Wasting by MUAC-HZ | 26.04 | 24.13 , 28.02 | 97.40 | 97.08 , 97.70 | 65.62 | 62.43 , 68.67 | 87.37 | 87.08 , 87.65 |
| Wasting by MUAC-UALG | 39.43 | 37.29 , 41.59 | 92.71 | 92.20 , 93.20 | 50.89 | 48.72 , 53.06 | 88.88 | 88.52 , 89.22 |

MUAC-125: mid-upper-arm circumference < 125 mm; MUAC-138: mid-upper-arm circumference < 138 mm; MUAC- HZ: MUAC-height Z score; MUAC-UALG: mid-upper-arm circumference by upper arm length group.

MUAC-125 had a lower PPV (57.87% [54.15; 61.50]) than MUAC-HZ (65.62% [62.43; 68.67]) and a lower NPV than that of all other indicators. MUAC-138 had the lowest PPV (35.42% [34.42; 36.44]) although the NPV was the highest among the indicators (92.89% [92.43; 93.32]).

## Discussion

In this study, we demonstrated two principal results related to the use of MUAC-UALG. First, the use of UAL along with MUAC enhanced WHZ based wasting prevalence estimation (table 2). Wasting prevalence evaluated by MUAC-UALG was the closest to that of WHZ < −2 when compared to other existing diagnosis methods. Using MUAC-125 and MUAC-HZ, wasting prevalence was three and two times lower than that of WHZ < −2 respectively. Wasting prevalence determined by MUAC-138 was almost three times higher than that of WHZ < −2. Fixed cut-off MUAC often overestimates or underestimates the number of wasting cases, depending on the threshold chosen.^9,13^ A fixed cut-off of 138 mm makes it possible to diagnose cases of wasting in older children but overestimates the number of wasting cases in the youngest children. Wasting prevalence according to the WHO standard MUAC cut-off of 125 mm is two times lower than that determined by the WHZ <-2 (table 2). When using a fixed cut-off at 138mm, the prevalence is two times higher than the prevalence using WHZ <-2. With the MUAC-UALG method, more wasted children belonging to UAL groups 2 and 3 can be diagnosed.

Second, the use of UAL in combination with MUAC enhanced the wasting diagnosis accuracy. We selected MUAC cut offs for each UALG in such a way to minimize the number of false positives (S1). Higher sensitivity could be obtained by selecting higher MUAC cut offs for each UALG, but we believe that this approach would have a negative impact on the malnutrition management system. Although the whole community needs nutrition interventions, those who are malnourished need it more. In our study, around 82 % (1-Sensitivity) of children with WHZ < −2 were not diagnosed with wasting when the current WHO MUAC cut off (MUAC-125) was used (table 3). The use of this single cut-off leaves older children behind, but using a higher single cut off is not adequate either. Indeed, an increasingly high rate of non-malnourished children could rise health workers’ burden and affect the quality wasting management. Although MUAC-ULAG alone could not detect all malnourished children, the overlap between WHZ and MUAC-UALG is higher than the overlap between MUAC-125 and WHZ. The MUAC-UALG method allows children’s age (for non-stunted children) and height to be taken into account through their arm length, unlike with the MUAC-125 approach. Fiorentino and colleagues showed that MUAC-125 was more adapted to younger children^(20)^. Thus, this method will allow field workers to diagnose more wasted children according to WHZ compared to the use of MUAC 125. MUAC by age group could be considered as a viable method but would not be accurate in stunted children. Moreover, children’s ages are not always easy to determine in the field, whereas UAL can be measured very easily. Fiorentino and colleagues had proposed different cut-offs according to age group and sex for children under five years old. With their method, the sensitivity ranged between 68% and 70% but the false positive rate was high, ranging between 30% and 32%^(20)^. Further studies on MUAC-UALG that evaluate the link with mortality are needed. Studies investigating wasting diagnostic methods could consider the MUAC-UALG as a diagnosis mean for comparison in the future^(21)^. Except for the MUAC-HZ for which ENA software did not provide values for 79 children, each indicator’s accuracy was calculated in the same population. Thus, indicator accuracies were compared with no risk of statistical bias. The MUAC-UALG method does not require any harmful nor stressful actions against children.

The study was conducted in the Mauritanian population which is not representative of the world population. However, a multi-centric study in different populations is feasible given the simplicity of collecting children’s UAL. WHZ was used as a reference standard for this study although this index is only a proxy for wasting. The overlap ratio between WHZ and MUAC varies by country.^5^ However, WHZ is widely used and accepted for wasting prevalence estimation around the world by the WHO. A more specific wasting diagnosis tool is needed in the future to compare with MUAC-UALG. Other alternative approaches could be used to evaluate the accuracy of MUAC-UALG method to identify more vulnerable children. Thus, MUAC-UALG mortality and or morbidity prediction capacity, and its association with wasting clinical biomarkers among children with low grade inflammation status could be considered.

At the community level, compared to the WHZ method, it is easier to use the MUAC-UALG which does not require any investment in equipment to measure height and weight. Measuring height and weight can be a challenge in emergency settings such as in corona virus disease 2019 (COVID-19) context. The portability of the MUAC tape is an advantage for its adoption by community health workers. The cost is also much lower than a scale measuring height and weight. Three MUAC tapes with different cut-offs according to UALG can be used by community health workers in the field for wasting diagnosis.

This study is aligned with the Council of Research & Technical Advice on Acute Malnutrition (CORTASAM) recommendations regarding the priority research^(22)^. Indeed, CORTASM group has recognized that the current MUAC admissions criteria for wasting (MUAC-115mm) does not select for all high-risk children, leaving behind some children who would be diagnosed as wasted by WHZ or WAZ methods. More research is needed concerning the options available to identify these high-risk children and ensure successful diagnosis and treatment, but the MUAC-UALG method is a promising candidate.

To our knowledge, the use of UAL in wasting diagnosis has never been proposed. This method does not add any additional tasks to the diagnostic process and has the potential to improve it. This method could be adopted in the field as a part of monitoring nutritional status of children and as an admission criterion in community-based management of acute malnutrition. Like MUAC-height or MUAC-age z-score, future studies aimed at the creation of a MUAC-UAL z-score should be considered. Using upper arm length-for-age z-score could also be considered as a substitute for the height-for-age method in diagnosing cases of chronic malnutrition. Indeed, UAL is simpler and less expensive than height measurement. A comparison of each child’s UAL with a same age and sex reference population could be considered for stunting diagnosis. Thus, in nutrition programs, weight-for-age monitoring could be supplemented with UAL-for-age in cases where children’s height is not known.

Beside wasting, obesity is also a major concern even in LMIC. Increasing the MUAC cut-off for wasting diagnosis for all children could have a negative impact if many non-wasted children who will receive treated. It could also prevent those in need to get enough supplements in an event of shortage. Our data showed that 9.6% of children were considered as wasted despite having a Body Mass Index Z-score > −1 when MUAC < 138mm. With MUAC-UALG this percentage drops to 2.6%.

## Conclusion

The diagnosis of wasting by a fixed cut-off MUAC has limitations that can be mitigated by the use of MUAC-for-height and MUAC-for-age indicators. The complexity of accurately collecting age and height in the field makes MUAC-UALG a good alternative for wasting diagnosis and prevalence estimation. MUAC-UALG could be used in emergency setting such as in COVID-19 context. The sensitivity and specificity of this method is higher than that of fixed MUAC cut-off methods and remains close to that of the MUAC-for-height and MUAC-for-age methods in Mauritanian children. Thus, using UAL along with MUAC enhances the accuracy of wasting diagnosis and the estimation of wasting prevalence. Future studies involving data from more children in different regions may lead to new perspectives on the use of MUAC-UALG as an anthropometric measure to diagnose wasting in developing countries. We recommend the inclusion of arm length in every national nutritional survey to collect more data for a multi-centric study.

## Data Availability

Data could be shared upon a reasonable request after publication.

## Acknowledgments

We would like to thank all participants and all investigators for their effort in data collecting.

## Notes

### Competing Interest Statement

One author, Mouhamed BARRO works now for Nutriset S.A.S France but this work started before he joined the company and is independent to his activity at the company.

### Clinical Trial

This study was not registered because it was not a real intervention study, since (except the upper arm length which was added), all data were collected from an annual routine nutritional survey without any protocol modification. A steering and ethics committee has been set up by the Ministry of Health and UNICEF to validate the protocol of the survey.

### Funding Statement

No author has received any specific funding for this study.

